# Penalized regression for left-truncated and right-censored survival data

**DOI:** 10.1101/2021.02.09.21251373

**Authors:** Sarah F. McGough, Devin Incerti, Svetlana Lyalina, Ryan Copping, Balasubramanian Narasimhan, Robert Tibshirani

## Abstract

High-dimensional data are becoming increasingly common in the medical field as large volumes of patient information are collected and processed by high-throughput screening, electronic health records (EHRs), and comprehensive genomic testing. Statistical models that attempt to study the effects of many predictors on survival typically implement feature selection or penalized methods to mitigate the undesirable consequences of overfitting. In some cases survival data is also left-truncated which can give rise to an immortal time bias, but penalized survival methods that adjust for left truncation are not commonly implemented. To address these challenges, we apply a penalized Cox proportional hazards model for left-truncated and right-censored survival data and assess implications of left truncation adjustment on bias and interpretation. We use simulation studies and a high-dimensional, real-world clinico-genomic database (CGDB) to highlight the pitfalls of failing to account for left truncation in survival modeling.

## 1 Introduction

The recent development and accessibility of genomic (DNA sequencing, microarrays), proteomic (immunohistology, tissue arrays), clinical (electronic health records), digital (wearables), and imaging (PET, fMRI) technologies have led to an increasing complexity and dimensionality of patient health information. The amount of person-level and disease-specific detail captured by these technologies offers an exciting new opportunity to understand patient history and prognosis by combining these molecular data with macro-scale clinical data on patient characteristics, treatment, and outcomes. Specifically, these data may offer insights into risk factors affecting patient-related outcomes leading to interest in developing prognostic tools to improve clinical decision-making and patient care ^1,2,3,4^. Given the hundreds or even thousands of molecular, patho-physiological, and clinical parameters that may be readily available for each patient, developing an appropriate statistical model that is robust to overfitting is critical to ensure good model performance and generalizability. Survival analysis is of particular interest given that overall survival is often a primary outcome of interest.

In a high-dimensional setting, computationally efficient regularization and shrinkage techniques have been developed ^5,6,7^ for the Cox model. Similar to ordinary least squares (OLS) regression, these penalized models estimate regression coefficients by minimizing the sum of squared errors, but place a constraint in the equation to minimize the sum of coefficients. This constraint (controlled by a parameter, *λ*) penalizes a high number of variables in the model and shrinks the coefficients of less influential features towards zero - acting, in cases when some coefficients are shrunk fully to zero, as automatic feature selection. Common penalized approaches include lasso ^8^, ridge ^9,10^, and elastic net ^11^, with extensions such as the adaptive lasso ^12^.

A common challenge in survival data is that patients are often included in the data only after having been at risk (e.g. diseased) for some time so that patients with shorter survival times are unobserved. In other words, the data are left-truncated because patients are only included if they survive beyond some initial milestone that marks entry into the observed data. An analysis of the observed patients is subject to an immortal time bias because observed patients cannot die prior to entering the study cohort ^13,14^. An example in medicine is a dataset describing patients who receive a specific diagnostic or genetic test; patients who die before having the chance to be tested are excluded from the data, and consequently their survival times do not contribute to overall survival estimates. Traditional survival models are inappropriate in this case and predicted survival probabilities are overestimated because of the exclusion of short survival times, biasing the study sample and consequently the estimation of the baseline risk and regression coefficients. Such bias has been observed in large oncology cohorts marked by a milestone entry, including patients who receive genomic tests ^15^ and specialized treatments ^16^. These databases are increasingly used to understand patient survival to support clinical decision-making, motivating the need to identify and address this survivor selection bias in practice.

Left truncation is a well known feature of medical data and there are methods to adjust for it ^17^. The Kaplan-Meier estimator of the survival function requires almost no modifications as it is only necessary to change the risk set so that individuals are only at risk of an event after study entry ^18^. Similarly, with left truncation, the likelihood function in a parametric model can be adjusted so that it is conditional on survival to the entry time.

Here, we apply a penalized Cox proportional hazards model for left-truncated and right-censored (LTRC) survival data and critically assess scenarios in which bias arises from and is exacerbated by left truncation. We apply the method to simulated data and a real-world use case from a de-identified clinico-genomic database (CGDB). Our results highlight the importance of adjusting for left truncation for both model interpretation and prediction. Furthermore, we show that use of inappropriate metrics for model evaluation can result in overly optimistic conclusions about the performance of predictive models. For implementation of the proposed method, we use the left truncation extension to survival modeling introduced in version 4.1 of the R package **glmnet** ^19^.

The remainder of this paper is organized as follows: In Section 2, we introduce the penalized regression and left truncation framework and present the method that incorporates both. Section 3 presents the simulation results and Section 4 considers a real-world dataset that combines electronic health records and genomic testing information. Finally, we discuss general takeaways and survival modeling recommendations in Section 5.

## 2 Methods

### 2.1 Data and notation

Consider a typical survival analysis framework in the absence of truncated survival times. For each patient *i*, we have data of the form *y*_*i*_, *e*_*i*_, *x*_*i*_ where *y*_*i*_ is the observed survival time, defined as the time between baseline and event, *e*_*i*_ is the event or failure of interest (e.g. death), and *x*_*i*_ = (*x*_*i*,1_, *x*_*i*,2_ …, *x*_*i,p*_) is a vector of predictors of length *p* used in the regression model. The event or failure of interest *e*_*i*_ is 1 if the event is observed or 0 if the event is right-censored. Suppose there are *m* unique event times, and let *t*_1_ *< t*_2_ *<* … *< t*_*m*_ be the ordered, unique event times. In this framework, all subjects are assumed to be at risk of the event starting at a commonly-defined “time zero” (a milestone that serves as a reference point for survival such as date of diagnosis) and observed over the interval [0, *T*] where *T* is the maximum follow-up time. Let *j*(*i*) be the index of the observation with an event at time *t*_*i*_. A Cox proportional hazards model is commonly used to model the relationship between survival times *y*_*i*_ and predictors *x*_*i*_ assuming a semi-parametric hazard of the form

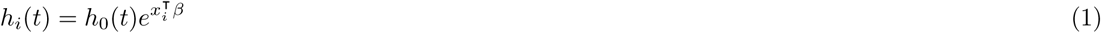

where *h*_*i*_(*t*) is the hazard for patient *i* at time *t, h*_0_(*t*) is the baseline hazard shared across subjects, and *β* is a length *p* vector of predictor coefficients.

Cox ^20^ proposed a partial likelihood for *β* without involving the baseline hazard *h*_0_(*t*)

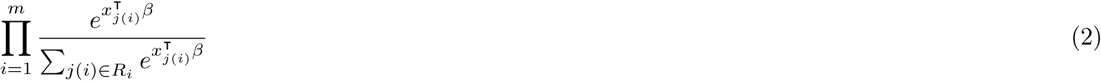

where *R*_*i*_ = *{j* : *y*_*j*_ *≥ t*_*i*_*}* denotes the set of indices *j*(*i*) who are “at risk” for failure at time *t*_*i*_, called the risk set. Maximizing the partial likelihood solves for *β* and allows for inferences on the relationship between survival time and the set of predictors.

### 2.2 Left truncation

In the absence of left truncation, the risk set *R*_*i*_ = *{j* : *y*_*j*_ *≥ t*_*i*_*}* assumes that every individual is at risk of an event at his or her respective time 0 and continues to be at risk until the event occurs or the individual is censored. In this sense, all individuals enter the risk set at the same time 0 and leave only at death or censoring. However, when survival times are left-truncated, individuals are truly at risk prior to entering the study cohort (e.g. prior to being observed for follow-up). Take Figure 1 as an example.

**Figure 1:**
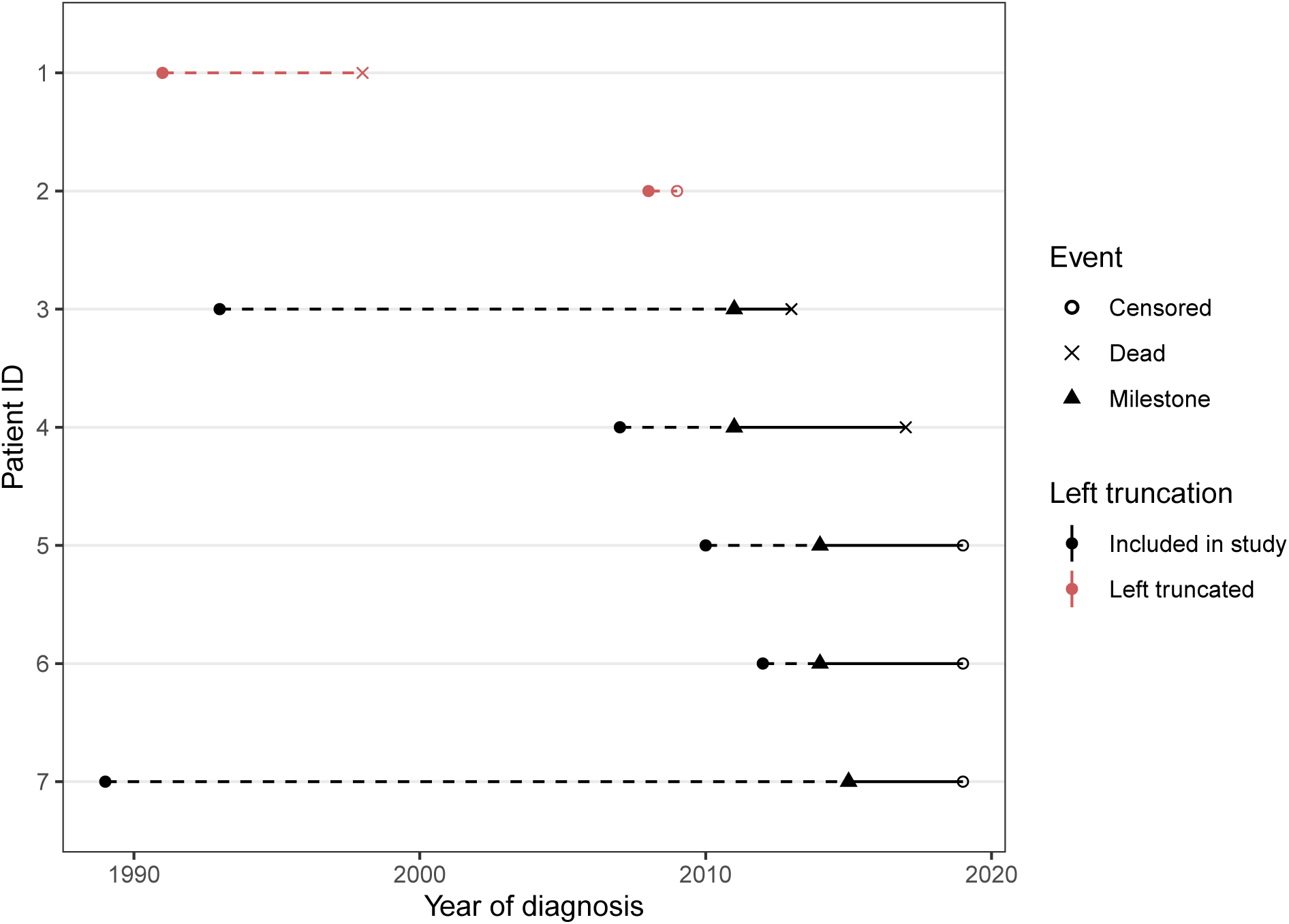
Left-truncated and right-censored patient follow-up in a hypothetical study cohort, ordered chronologically by event time. Patients who receive a diagnosis (closed circle) become eligible to enter the cohort after reaching a milestone (black triangle), for example a genomic test. Patients are followed until death (open circle) or censoring (cross). However, patients who die before reaching the milestone are left-truncated (in red), and only those who have survived until eligibility (in black) are observed. Left truncation time, or the time between diagnosis and cohort entry, is shown with a dashed line.

Survival is defined as the time from diagnosis until death, but patients are not eligible to enter the cohort until a milestone is reached (black triangle). Patients 1 and 2 (in red) are excluded from the analysis because of death and right censoring, respectively, prior to study eligibility. A naive analysis on this data would therefore be biased because it only contains survival time for “super-survivors” (those who have survived a long time, such as patients 3 and 7) and the recently-diagnosed.

Fortunately, the partial likelihood can be easily modified to make *R*_*i*_ the set of individuals who are at risk (have entered the study and are not censored or failed) at that time. Let individual *i* have survival data of the form (*v*_*i*_, *y*_*i*_, *e*_*i*_, *x*_*i*_), where individual *i* enters the study at time *v*_*i*_. The left-truncated risk set *R*_*i\**_ = *{j* : *y*_*j*_ *≥ t*_*i*_ *> v*_*j*_*}*. In other words, subjects are only considered at risk at a given event time after they have been observed to be at risk (i.e. exceeded their left truncation time, shown with a solid black line in Figure 1). For example, at the time of death of Patient 3 in Figure 1, only Patient 4 has reached the milestone for study entry and is at risk. Patients 5, 6, and 7 reach their milestones at later time points.

The partial likelihood in Equation 2 can then be evaluated by substituting *R*_*i\**_ to make inferences on *β* in the case of left truncation.

### 2.3 Penalized regression with left-truncated and right-censored data

Penalized regression methods such as *l*_1_ (lasso) ^8^ and *l*_2_ (ridge) ^21^ regression, elastic net regression ^11^, and SCAD (Smoothly Clipped Absolute Deviation) ^22^ have been developed to overcome the limitation of fitting a model to a large number of predictors. In a high-dimensional feature space, where the number of predictors *p* is large relative to the sample size *n*, a model may exhibit low bias but suffer from poor prediction accuracy and limited generalizability, a consequence of overfitting the data. Penalized approaches, which add a regularization penalty to constrain the size of the *β*, reduce the complexity of the model and mitigate the negative effects of overfitting. Note that in high-dimensional settings where the number of predictors is larger than the sample size (i.e., *p* > *n*), a standard regression cannot be fit at all.

The extension of penalized regression methods to survival data have been extensively described and applied, for example in microarray gene expression data ^1^, transcriptomes ^23^, and injury scale codes ^24^. Like linear regression, the performance and fit of a Cox proportional hazards model is known to deteriorate with large *p*, providing an advantage to penalized approaches in high-dimensional, time-to-event feature sets.

Simon et. al ^7^ introduced a fast, pathwise algorithm to fit a regularized Cox proportional hazards model solving for

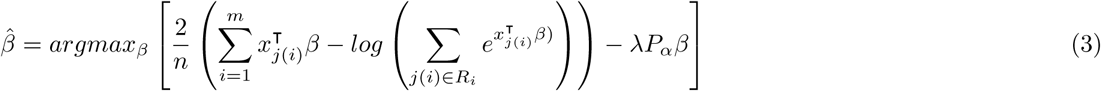

where *β* is the vector of regression coefficients and is maximized over Equation 3 subject to a constraint defined by the penalty *λ · P*_*α*_*β*. As described elsewhere ^7^, the penalty can take the form of the *l*_1_ (lasso), *l*_2_ (ridge), or elastic net penalty, and the algorithm fits over a path of *λ* values via cyclical coordinate descent. Instead of solving a least squares problem at each iteration, the algorithm solves a penalized reweighted least squares problem, in which observation weights *w* are defined in the coordinate descent least squares minimization step. The optimal *λ* can be identified through cross-validation. Tied survival times are handled via the Breslow approximation ^25^ and details provided in ^7^.

Applying the standard penalized Cox implementation in the software **glmnet** to left-truncated survival data will result in biased estimates. However, an extension to LTRC data is straightforward. We solve for Equation 3 with a modification to the risk set *R*_*i*_:

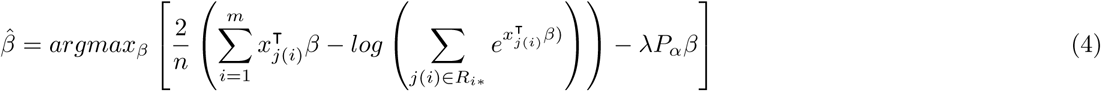

where the left-truncated risk set *R*_*i\**_ = *{j* : *y*_*j*_ *≥ t*_*i*_ *> v*_*j*_*}*; that is, subjects are only considered at risk after their left truncation times. This implementation is available in the release of **glmnet** version 4.1 ^19^.

### 2.4 Real-world use case

We consider the Clinico-Genomic Database offered jointly by Flatiron Health and Foundation Medicine (FH-FMI CGDB) ^26^. The CGDB is a US-based de-identified oncology database that combines real-world, patient-level clinical data and outcomes with patient-level genomic data to provide a comprehensive patient and tumor profile. The de-identified data originated from approximately 280 US cancer clinics (800 sites of care). Patients in the CGDB have received at least one Foundation Medicine comprehensive genomic profiling test as well as have had their electronic health records captured by Flatiron Health. The retrospective, longitudinal EHR-derived database comprises patient-level structured and unstructured data, curated via technology-enabled abstraction, and were linked to Foundation Medicine genomic data by de-identified, deterministic matching. Genomic alterations were identified via comprehensive genomic profiling (CGP) of >300 cancer-related genes on FMI’s next-generation sequencing (NGS) based FoundationOne panel ^27,28^. To date, over 400,000 samples have been sequenced from patients across the US. The data are de-identified and subject to obligations to prevent re-identification and protect patient confidentiality. Altogether, the CGDB represents thousands of potential predictors per patient, ranging from demographic, laboratory, and treatment information to the specific mutations detected on each biomarker included in a Foundation Medicine baitset. Institutional Review Board approval of the study protocol was obtained prior to study conduct, and included a waiver of informed consent.

We take a subset of 4, 429 patients with stage IV, non-small cell lung cancer (NSCLC) in the CGDB as a use case for this study. The outcome of interest is survival time predicted from the date of stage IV diagnosis. Survival times are left-truncated because patients who die before receiving a Foundation Medicine test are logically not included in the cohort of Foundation Medicine test recipients.

As shown in Figure 2, the distribution of left truncation times (time from diagnosis to genomic testing) is right-tailed with approximately 21.5% of patients diagnosed more than one year prior to receiving the Foundation Medicine genomic test. Moreover, the correlation between diagnosis year and left truncation time is strongly negative (*ρ* = *−*0.63), suggesting the presence of left truncation survivor bias as patients with records in the CGDB diagnosed longer ago needed to survive more years before the availability of genomic testing.

**Figure 2:**
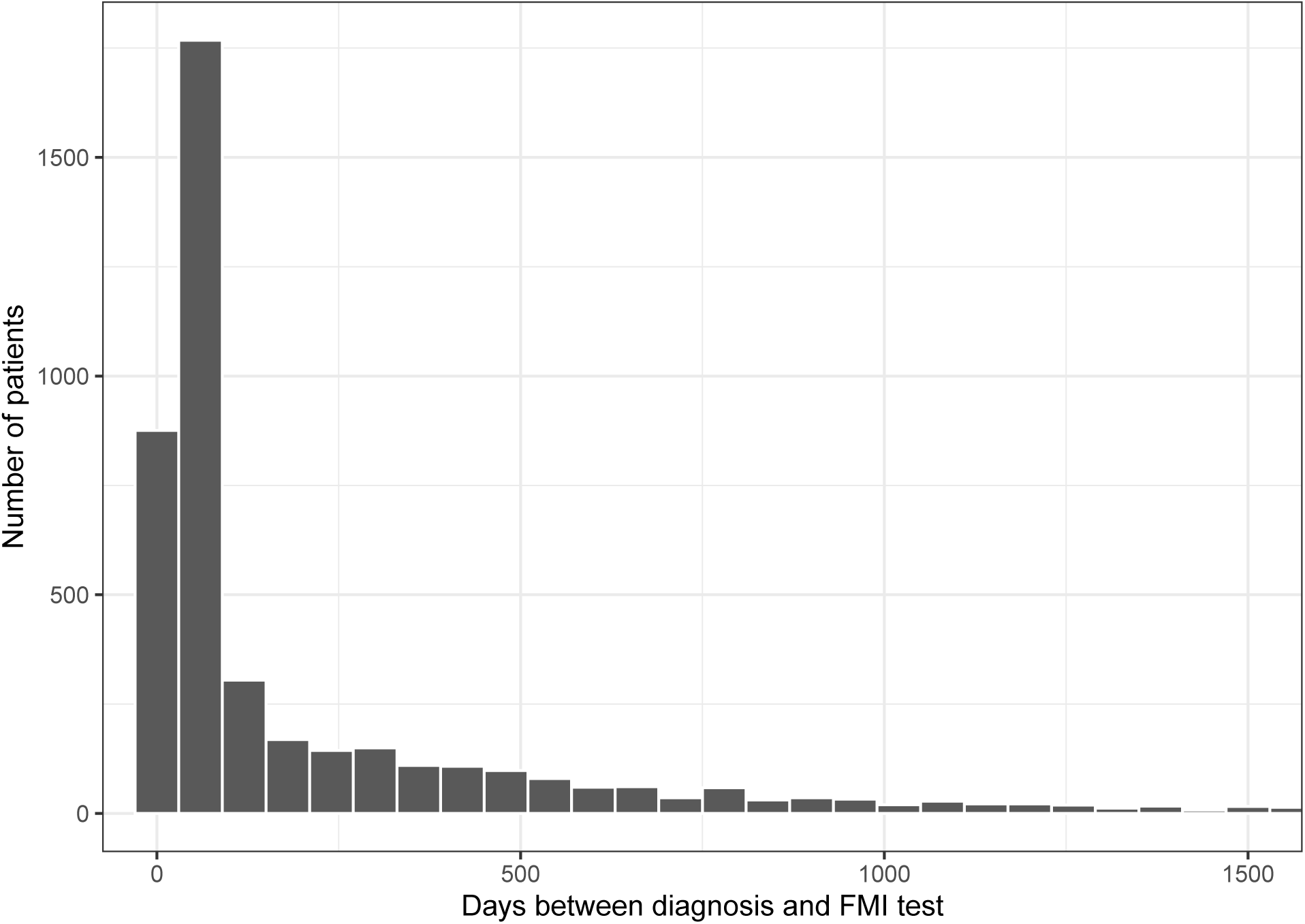
Distribution of left truncation time (days) in non-small cell lung cancer patients in the CGDB

Next, we use these data as the basis for a simulation study and a real-world application.

## 3 Simulation study

Simulations were conducted to assess the performance of the method with LTRC data. To set notation, let *T*_*i*_, *U*_*i*_, and *V*_*i*_ be latent survival, right censoring, and study entry times for patient *i*, respectively. The simulation proceeds by simulating *T*_*i*_, *U*_*i*_, and *V*_*i*_ from suitable survival distributions and assumes that *T*_*i*_, *U*_*i*_, and *V*_*i*_ are uncorrelated. The observed survival time is given by *Y*_*i*_ = min(*T*_*i*_, *U*_*i*_) and a patient is right-censored if *T*_*i*_ *> U*_*i*_. Patients are left-truncated (i.e., hidden from the sample) if *V*_*i*_ *> Y*_*i*_.

### 3.1 Data generating process

We generated *n* patients and *p* predictors to construct a *n × p* matrix, *X*. The first 11 predictors were from the CGDB and the remaining 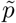 *p*variables were simulated binary variables to represent a comprehensive set of CGDB binary alterations. The CGDB variables were the same 11 variables used in the small *p* model in Section 4 and generated by randomly sampling rows of the original data with replacement. The matrix of binary variables, 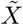 was simulated using a (latent) multivariate probit approach

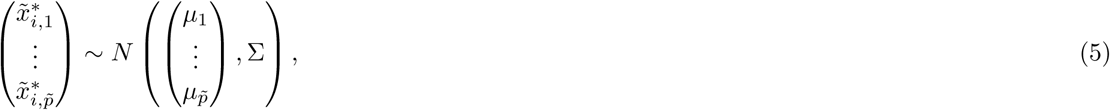

with the *k*th binary variable 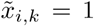 if 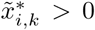 and 0 otherwise. Each diagonal element of Σ equals 1 and non-diagonal elements determine the correlation between variables. The probability, *π*_*k*_ that variable *k* was equal to 1 was randomly sampled from a uniform distribution ranging from .2 to .8; that is, we sampled *π*_*k*_ *∼ U* (0.2, 0.8) and set *µ*_*k*_ = Φ^*−*1^(*π*_*k*_) where Φ(*·*) is the cumulative density function of the normal distribution. Σ was generated by filling a 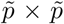 matrix, 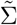, with *N* (0, 1) random variables and setting Σ = *S/D*^*T*^ *D* with 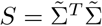 and *D* a vector containing the diagonal elements of *S*.

Latent survival time was simulated from a proportional hazards Weibull distribution with shape parameter *a* and scale parameter *m*_*i*_ based on evidence from the CGDB as detailed in Section 3.2. Predictors predict latent survival time through their effect on the scale parameter,

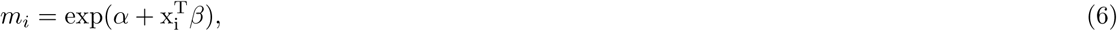

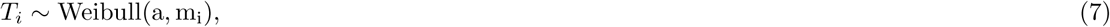

where *α* is an intercept and *β* is a vector of coefficients. *α* and values of *β* for the CGDB variables were estimated from the CGDB data. The probability that the coefficient of a binary variables was non-zero was 0.5 and drawn from a Bernoulli distribution. Non-zero coefficients were drawn from a uniform distribution with lower bound of *−*0.25 and upper bound of 0.25 so that the minimum and maximum hazard ratios were 0.78 and 1.28, respectively.

Time to right censoring was also sampled from a Weibull distribution but did not depend on predictors,

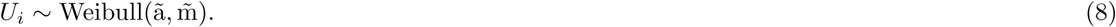

Lastly, a two-part model was used to simulate time to study entry so that a proportion 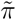 of patients could enter the study at *t* = 0 while a proportion 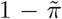 would enter at *t >* 0. The probability of an entry time greater than 0 was drawn from a Bernoulli distribution and positive entry dates were modeled using a lognormal distribution.

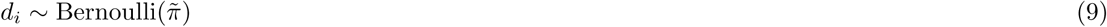

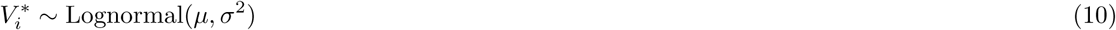

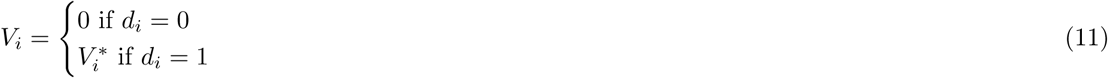

### 3.2 Calibration

The simulation was calibrated to be consistent with the CGDB NSCLC data. The Nelson-Aalen estimator of the cumulative hazard of overall survival was generally monotonically decreasing, which suggested that a Weibull distribution could adequately capture the baseline hazard. This was further supported by the similarity in fit between a spline-based survival model with one internal knot at the median of uncensored survival time and the Weibull model (Figure S1). As such, an intercept only Weibull model was used to estimate *a* and *α* in the simulation. Similarly, a Weibull distribution was used to model time to right censoring by reversing the censoring indicator in the model for overall survival. Both the model for overall survival and right censoring adjusted for left truncation.

The CGDB variables were standardized to have mean 0 and standard deviation 1 and coefficients were estimated using a Cox proportional hazards model. The full coefficient vector combined the coefficients from the CGDB variables with the coefficients from the simulated binary variables. The scale parameter was set to *m* = *α* + *x*^*T*^ *β − E*(*x*^*T*^ *β*) so that predicted survival averaged across patients was equal to predicted survival from the intercept only Weibull model.

Latent time to study entry is inherently unobserved, but we believe a two-part model can characterize the large fraction of tests in the observed data that occur near the diagnosis date (Figure 2). The probability of a positive entry time was set equal to 0.2 and the lognormal distribution for positive entry times was derived using a right skewed distribution with a median value of 1 and a mean value of 1.6, both measured in years. We found that this parameterization generated median survival times in the simulation that were similar to median survival in Kaplan-Meier estimates that did not adjust for left truncation in the CGDB (Figure S2).

### 3.3 Implementation

Datasets were simulated where each row contained information on a patient’s observed survival time (*y*_*i*_), a censoring indicator (*e*_*i*_), the time of study entry (*v*_*i*_). 75% of patients were assigned to a training set and the remaining 25% were assigned to a test set.

Within the training set, Cox models with lasso penalties were fit among the “observed” (i.e., non-truncated) patients. Cross-validation with 10 folds was used to tune the model and the value of *λ* that minimized the partial likelihood deviance was selected. Models were fit with and without adjusting for left truncation. All models adjusted for right censoring.

Predictions and model evaluations were performed on the test set. The models were evaluated using the “complete” sample consisting of both truncated and non-truncated patients. The use of the complete sample for model evaluation is a key advantage of a simulation study since it is unobservable in real-world applications.

This process was repeated 200 times. For each iteration, models were evaluated based on a dataset consisting of *n* = 5, 000 patients. Simulations were performed in both small and large *p* scenarios. In the small *p* scenario, 10 binary variables were simulated in addition to the 11 CGDB variables so that *p* = 21; in the large *p* scenario, 1, 000 binary variables were simulated so that *p* = 1, 011.

### 3.4 Results

We used calibration curves ^29^ to compare models with and without a left truncation adjustment. Each point in the plot consists of simulated patients in a given decile of predicted survival at a given time point. The x-axis is the averaged predicted survival probability and the y-axis is the pseudo observed survival probability computed using the Kaplan-Meier estimator. Perfectly calibrated predictions are those that lie along the 45 degree line so that predicted and observed survival probabilities are equal. The plots consist of patients in the complete sample and are therefore a good test of the effectiveness of the left truncation adjustment.

Calibration curves are displayed in Figure 3 for both the small and large *p* scenarios. Predicted survival probabilities from the model that does not adjust for left truncation are too high because the model is fit on the observed sample consisting only of patients that survived long enough to enter the sample. In other words, failing to adjust for left truncation results in an overestimation of survival probabilities. In contrast, the left-truncated adjusted model corrects for this bias. The small *p* model is almost perfectly calibrated with points along the 45 degree line.

**Figure 3:**
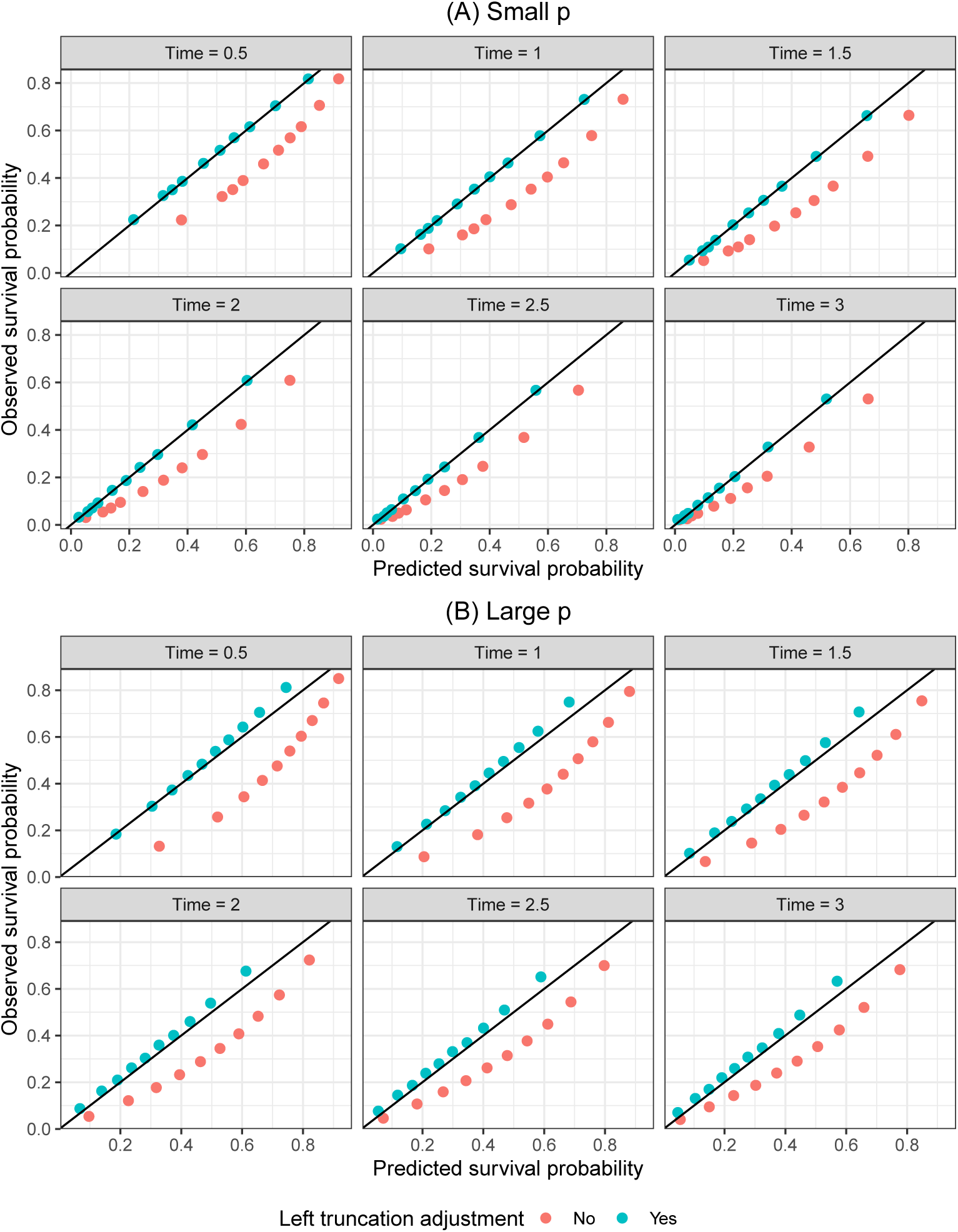
Calibration of survival predictions for lasso model in simulation. Notes: The Cox model with lasso penalty using the training data and was subsequently used to predict the survival function for each patient in the test set. The small and large *p* models contained 21 and 1, 011 predictors, respectively. Patients were divided into deciles at each time point based on their predicted survival probabilities. Each point in the plot represents patients within a decile. The “Predicted survival probability” is the average of the predicted survival probabilities from the Cox model across patients within each decile and the “Observed survival probability” is the Kaplan-Meier estimate of the proportion surviving within each decile. A perfect prediction lies on the black 45 degree line.

The results of the large *p* scenario are similar, although calibration is more difficult than in the lower dimensional setting with points lying farther from the 45 degree line for patients with higher survival probabilities. Predictions tend to be more accurate for patients with predicted survival close to and below 0.5, but slightly underestimated for patients with higher survival probabilities. Probability rescaling has been effective in such cases, for example with the use of Platt scaling ^30^ or isotonic regression ^31^, though implementation is less straightforward in a survival context^32^.

A common metric used to evaluate survival models is the C-index ^33^, which provides a measure of how well a model will discriminate prognosis between patients. We computed the C-index using predictions from the Cox model with lasso penalty on high-dimensional data. The results are presented in Table 1 and suggest that care should be taken when using the C-index to evaluate survival models in the presence of left-truncated data. When using the observed sample to evaluate the model, the C-index is higher in a model that does not adjust for left truncation, even though it is poorly calibrated. Furthermore, even when using the complete sample, the C-indices are very similar.

**Table 1:** Comparison of model discrimination in the simulation.

| Left truncation adjustment | Test sample | C-index |
| --- | --- | --- |
| No | Observed | 0.70 |
| Yes | Observed | 0.66 |
| No | Complete | 0.65 |
| Yes | Complete | 0.65 |

Cox models with lasso penalty were fit using the high-dimensional simulated data.

These results are perhaps not surprising. For example, when computing the C-index in the observed sample, the model is evaluated in a test set that suffers from the same bias as the training set. Moreover, the C-indices may be nearly identical in the complete sample because we did not simulate a dependence between the predictors and entry time. In other words, the bias in the coefficients is based only on the indirect correlation between the predictors and left truncation induced by (i) the correlation between the predictors and survival and (ii) the correlation between survival and the probability a patient is left-truncated. As we will see in Section 4, the impact of left truncation adjustment on the estimated values of the coefficients (and by extension the C-index) can be considerably larger if there is a more direct correlation.

## 4 Real-world data application: Patients with non-small cell lung cancer in the Clinico-Genomic Database (CGDB)

We fit models using 4, 429 patients with stage IV, NSCLC. 75% of patients were assigned to a training set and the remaining 25% were assigned to a test set. The training set was then used to train and tune both small *p* and large *p* models with 11 and 639 predictor variables, respectively. Both un-penalized and lasso penalized Cox models were fit, with and without a left truncation adjustment. In the penalized models, the optimal value of the shrinkage parameter, *λ*, was selected using the value that minimized the partial likelihood deviance in 10-fold cross-validation. The models were evaluated by assessing predictions on the test set.

Both the small and large *p* models were designed to highlight left truncation in a real-world dataset and its impact on models, as opposed to optimizing predictive performance and the prognostic value of predictors. For this reason, model specifications were relatively simple, e.g. modeling linear and additive effects, applying no transformations to predictors (for instance, modeling mutations as binary), and including a variable, year of diagnosis, that is highly correlated with left truncation time but not necessarily with survival (Figure 2). Survival was predicted from diagnosis. For an evaluation of meaningful prognostic clinical and genomic factors in NSCLC, see for example Lai and colleagues ^34^.

The smaller model consisted of the following variables: year of diagnosis, age at diagnosis, race, practice type (academic or community center), and the mutational status (mutated/not mutated) of 3 known prognostic biomarkers for NSCLC (TP53, KRAS, and EGFR). Short variants (SV) were considered for each biomarker while copy number alterations (CN) and rearrangements (RE) were also considered for TP53. 3, 022 patients remained in the training set after dropping patients with missing values on at least one covariate.

The larger model included all non-biomarker variables from the smaller model and added nearly all mutations tested by Foundation Medicine. As the data come from both historic and current clinico-genomic testing products, the set of tested genes has changed over time. To resolve the resultant missing data problem, we constrained the mutation data to biomarkers with less than 30% missingness and imputed the remaining mutation data using k nearest neighbors ^35^, with k = 20. Biomarkers with zero variance or that were present in less than 0.1% of patients were also excluded.

Figure 4 displays hazard ratios for each model. The plot for the large *p* model only contains coefficients ranked in the top 10 by absolute value of the hazard ratio in either the left truncation adjusted or non-adjusted model. The arrows denote the impact of adjusting for left truncation on the hazard ratio. In some cases the adjustment can have a considerable impact on the hazard ratios, often moving a large hazard ratio all the way to 1 (i.e., to a coefficient of 0). This is most notable with diagnosis year, which has a hazard ratio of approximately 1.5 in both the small and large *p* models but is set to 1 after the left truncation adjustment. This effect is likely due to the strong negative correlation between calendar time and the probability that a patient is left-truncated.

**Figure 4:**
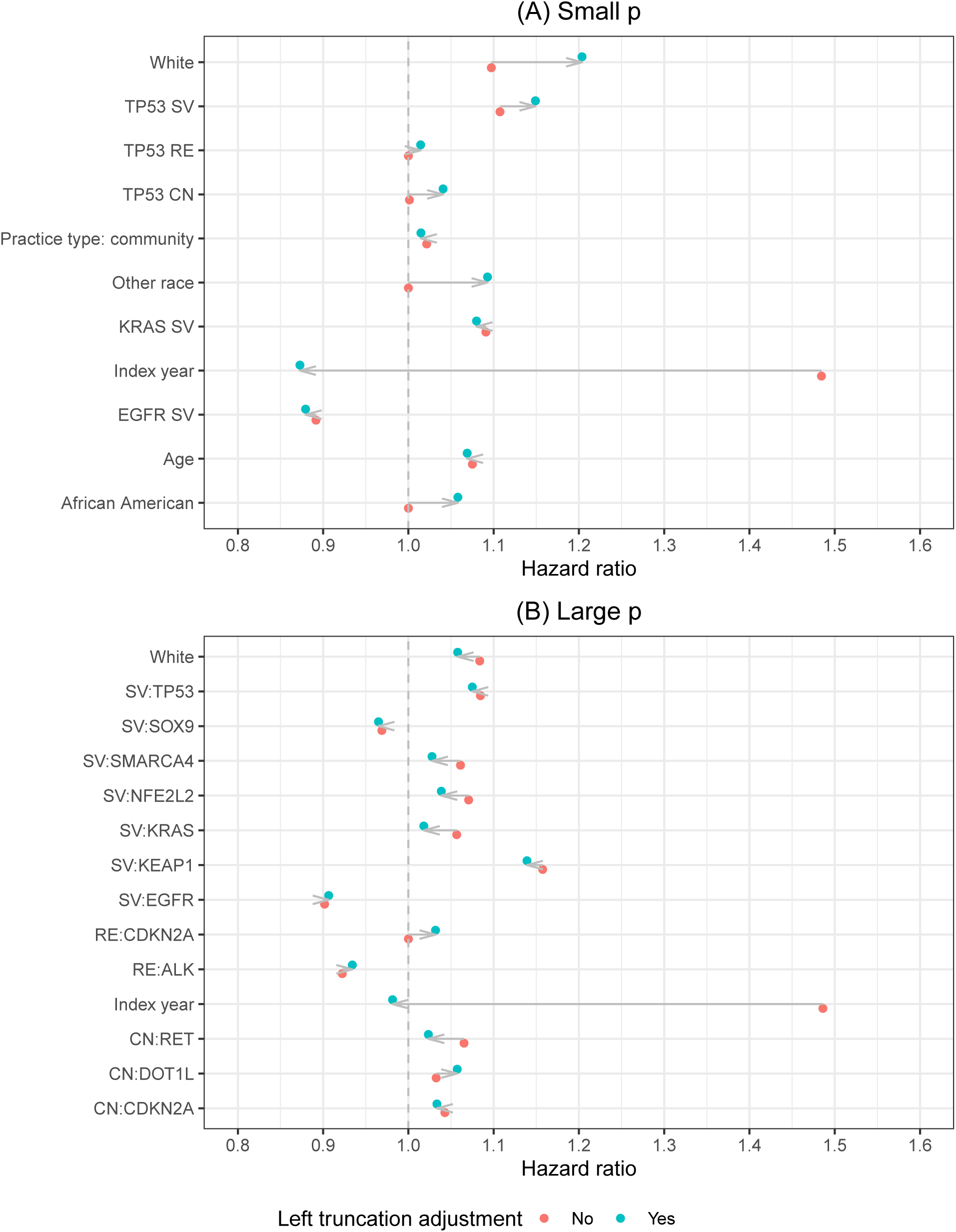
Hazard ratios from Cox lasso model. Notes: The figure for the “Large” *p* model only includes variables ranked in the top 10 by the absolute value of the hazard ratio in either the left truncation adjusted or non-adjusted model.

Table 2 and Figure 5 evaluate the models based on predictions on the test set. Table 2 compares models using the C-index. The C-index is relatively low in all of these illustrative models, though is artificially higher in the unadjusted models that do not account for left truncation—sometimes considerably so—and decreases after adjustment as the predictor hazard ratios are pushed toward 1 and there are fewer differences in predicted relative risks across patients. As expected, the lasso model has a higher C-index in the larger *p* setting as the Cox model is considerably overfitted.

**Table 2:**
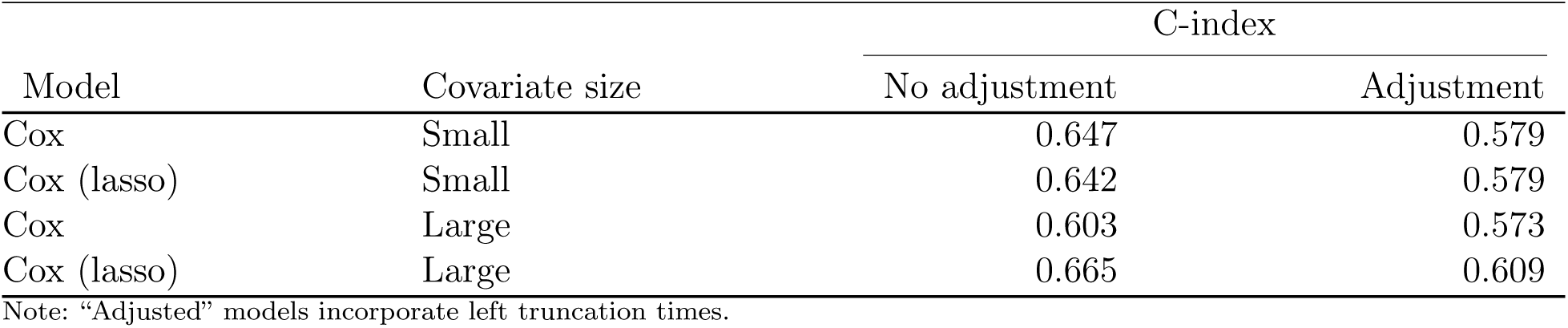
Comparison of model discrimination in the CGDB.

**Figure 5:**
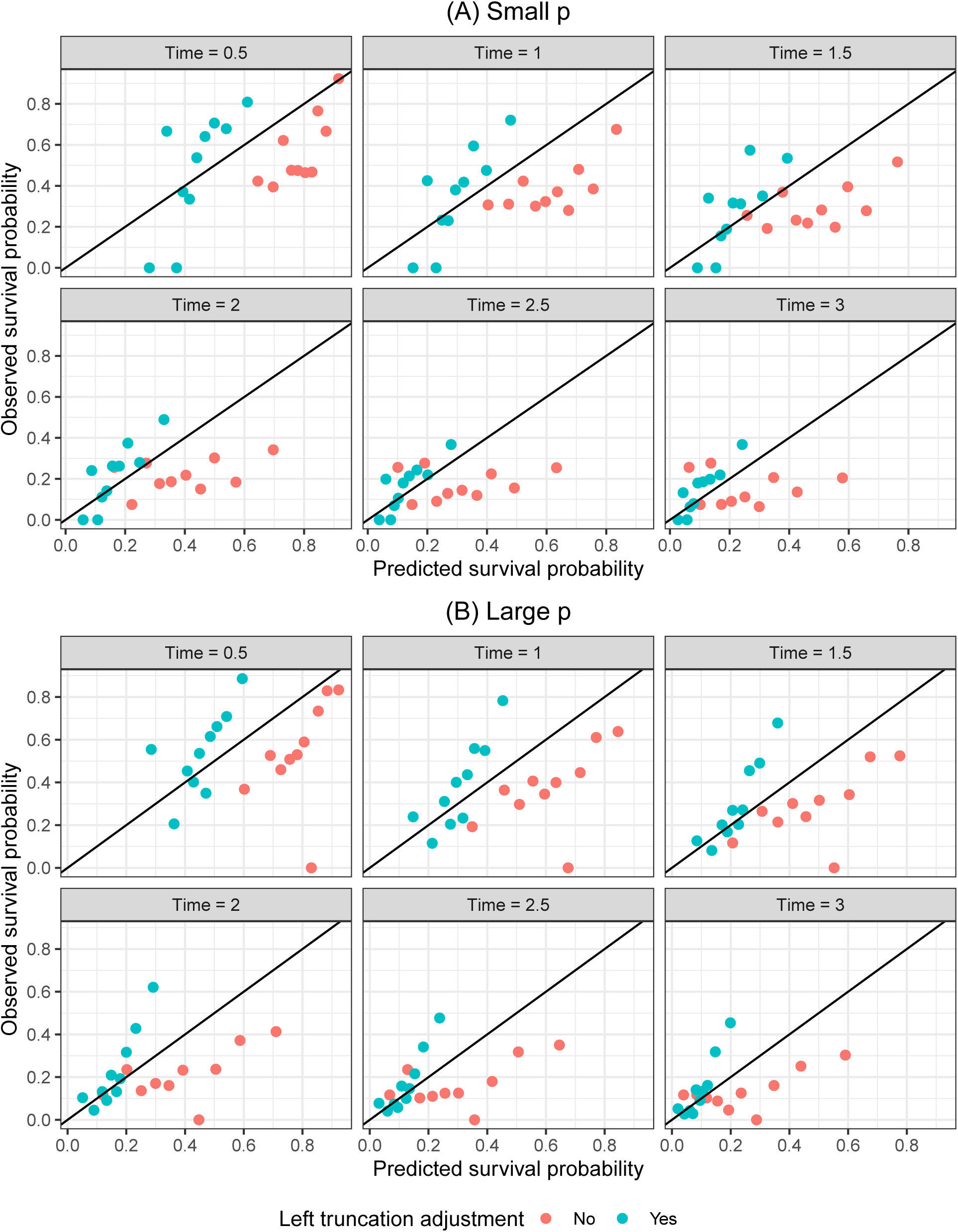
Calibration of survival predictions in the CGDB from the Cox lasso model

Figure 5 displays the same survival calibration curves presented in the simulation section. As in the simulation study, the unadjusted models overpredict survival probabilities; however, in contrast to the simulations, the adjusted models are poorly calibrated.

The adjusted models tend to be better calibrated for patients with the lowest survival probabilities but poorly calibrated for patients with higher survival probabilities. One possible reason for this result is that the adjusted models pushed many of the hazard ratios to 1 and do not adequately separate low and high risk patients. Probability rescaling such as Platt scaling or isotonic regression may be effective on the adjusted probabilities here, which resemble the characteristic sigmoid-shaped calibration curves produced by different learning algorithms, including the lasso.

## 5 Discussion

In this paper, we applied an approach for estimating penalized Cox proportional hazards model with LTRC survival data and compared it to penalized models designed for survival data that is right-censored but not left-truncated. Using simulation studies and examples from real-world EHR and genomics data, we showed that there is a need for approaches that can both adjust for left truncation and model high-dimensional data. In particular, our simulation shows that predictions from models that fail to adjust for left truncation will overestimate true survival probabilities whereas models that properly adjust can yield well calibrated survival predictions, even with high-dimensional data. Moreover, in our real-world use case, there were significant differences in coefficient estimates between models that did and did not adjust for left truncation.

Our results have important implications for specification, interpretation, and evaluation of prognostic survival models. Much of the difficulty arises because training and test sets are both subject to the same selection bias. In other words, a careful understanding of the data generating process is crucial and naive approaches can lead analysts astray. For example, our simulations suggest that the commonly used C-index cannot differentiate between models that mimic the true data generating process and those that do not, and may therefore lead to misleading conclusions in the presence of LTRC data. On the other hand, metrics that focus on predicted survival probabilities such as calibration curves are better able to determine whether a model can accurately predict the risk of an event.

In our real-world use case, an analyst could have mistakenly concluded that the high C-index in the non-adjusted model was indicative of good model performance when it was in fact driven primarily by a high correlation between predictors and left truncation. For instance, in our large *p* model the hazard ratio for the predictor “year of diagnosis” moved from being the largest in absolute value to 1 after adjusting for left truncation. The high C-index in the non-adjusted model was artificial because patients diagnosed earlier appeared to survive longer due to immortal time bias. To facilitate model interpretation, we recommend that models should be evaluated using both discrimination and calibration based metrics.

Although not the primary focus of this paper, our approach generated findings that were both consistent with prior literature in genomics and clinically plausible. One of the mutation features that had the highest importance was the presence of short variants in the gene KEAP1. This finding is supported by similar reports of risk-conferring status in an independent lung cancer cohort ^36^. Conversely, variants in EGFR and ALK were found to be protective in the CGDB data, putatively because mutations in these genes qualify patients to receive targeted therapies and therefore benefit from improved response and longer survival.

When making predictions with survival data is of interest, it is critical to define the risk set appropriately, as our work demonstrates. To do this requires recognizing the underlying process by which individuals are captured in the data in the first place; i.e., when and how individuals came to be included in the dataset of interest. One of the motivations for the current work derives from the advent of biobank and genomic test databases that store information on patient illness - presumably after the patient has been diagnosed and “at risk” for some time. In these data, overlooking the “delayed entry” of at-risk patients could result in misleading conclusions, as we demonstrate in this work. Additionally, patient cohorts marked by a milestone entry, such as genomic testing, may not only be subject to immortal time bias but also to a temporal selection bias. The latter may occur if left truncation time is correlated with survival time, for example, if genomic testing is ordered selectively for patients with worse prognosis. This was uncovered in a cohort of multi-stage cancer patients who received genomic testing ^15^ and can affect inference even in the presence of a left truncation adjustment. While the cohort of lung cancer patients in our real-world use case was restricted to stage IV diagnoses, mitigating this potential selection bias ^15^, it can be adjusted for by modeling the association between left truncation time and survival ^37,38^.

## Data Availability

The data that support the findings of this study have been originated by Flatiron Health, Inc. and Foundation Medicine, Inc. These de-identified data may be made available upon request, and are subject to a license agreement with Flatiron Health and Foundation Medicine; interested researchers should contact to determine licensing terms.
Code used to produce the analysis is publicly available at https://github.com/phcanalytics/coxnet-ltrc.

## Acknowledgments

We thank T. Hastie and K. Tay for their work in implementing left truncation-adjusted Cox models in **glmnet**. Competing interests: R. Tibshirani and B. Narasimhan are paid consultants for Roche.

## Data availability statement

The data that support the findings of this study have been originated by Flatiron Health, Inc. and Foundation Medicine, Inc. These de-identified data may be made available upon request, and are subject to a license agreement with Flatiron Health and Foundation Medicine; interested researchers should contact to determine licensing terms.

Code used to produce the analysis is publicly available at https://github.com/phcanalytics/coxnetltrc.

## References

1. Gui J, Li H. Penalized Cox regression analysis in the high-dimensional and low-sample size settings, with applications to microarray gene expression data. Bioinformatics 2005; 21(13): 3001–3008.

2. Wishart GC, Azzato EM, Greenberg DC, et al. PREDICT: a new UK prognostic model that predicts survival following surgery for invasive breast cancer. Breast Cancer Research 2010; 12(1): R1.

3. Ow GS, Kuznetsov VA. Big genomics and clinical data analytics strategies for precision cancer prognosis. Scientific reports 2016; 6: 36493.

4. Yousefi S, Amrollahi F, Amgad M, et al. Predicting clinical outcomes from large scale cancer genomic profiles with deep survival models. Scientific reports 2017; 7(1): 1–11.

5. Tibshirani R. The lasso method for variable selection in the Cox model. Statistics in medicine 1997; 16(4): 385–395.

6. Friedman J, Hastie T, Tibshirani R. Regularization paths for generalized linear models via coordinate descent. Journal of statistical software 2010; 33(1): 1.

7. Simon N, Friedman J, Hastie T, Tibshirani R. Regularization paths for Cox’s proportional hazards model via coordinate descent. Journal of statistical software 2011; 39(5): 1.

8. Tibshirani R. Regression shrinkage and selection via the lasso. Journal of the Royal Statistical Society: Series B (Methodological) 1996; 58(1): 267–288.

9. Hoerl AE, Kennard RW. Ridge regression: applications to nonorthogonal problems. Technometrics 1970; 12(1): 69–82.

10. Hoerl AE, Kennard RW. Ridge regression: Biased estimation for nonorthogonal problems. Technometrics 1970; 12(1): 55–67.

11. Zou H, Hastie T. Regularization and variable selection via the elastic net. Journal of the royal statistical society: series B (statistical methodology) 2005; 67(2): 301–320.

12. Zou H. The adaptive lasso and its oracle properties. Journal of the American statistical association 2006; 101(476): 1418–1429.

13. Lévesque LE, Hanley JA, Kezouh A, Suissa S. Problem of immortal time bias in cohort studies: example using statins for preventing progression of diabetes. Bmj 2010; 340: b5087.

14. Giobbie-Hurder A, Gelber RD, Regan MM. Challenges of guarantee-time bias. Journal of clinical oncology 2013; 31(23): 2963.

15. Kehl KL, Schrag D, Hassett MJ, Uno H. Assessment of temporal selection bias in genomic testing in a cohort of patients with cancer. JAMA Network Open 2020; 3(6).

16. Newman NB, Brett CL, Kluwe CA, et al. Immortal time bias in national cancer database studies. International Journal of Radiation Oncology*Biology*Physics 2020; 106(1): 5–12.

17. Kalbfleisch JD, Prentice RL. The statistical analysis of failure time data. 360. John Wiley & Sons. 2011.

18. Tsai WY, Jewell NP, Wang MC. A note on the product-limit estimator under right censoring and left truncation. Biometrika 1987; 74(4): 883–886.

19. Friedman J, Hastie T, Tibshirani R, et al. glmnet: Lasso and Elastic-Net Regularized Generalized Linear Models. 2021. R package version 4.1.

20. Cox DR. Regression models and life-tables. Journal of the Royal Statistical Society: Series B (Methodological) 1972; 34(2): 187–202.

21. Tikhonov AN. Solution of incorrectly formulated problems and the regularization method. Soviet Mathematics 1963; 4: 1035–1038.

22. Xie H, Huang J. SCAD-penalized regression in high-dimensional partially linear models. Annals of Statistics 2009; 37(2): 673–696.

23. Wu TT, Gong H, Clarke EM. A transcriptome analysis by lasso penalized Cox regression for pancreatic cancer survival. Journal of Bioinformatics and Computational Biology 2011; 9(1): 63–73.

24. Mittal S, Madigan D, Cheng J, Burdc R. Large-scale parametric survival analysis. Statistics in Medicine 2013; 32(23): 3955–3971.

25. Breslow NE. Contribution to the Discussion of the Paper by D.R. Cox. Journal of the Royal Statistical Society: Series B (Methodological) 1972; 34(2): 216–217.

26. Singal G, Miller PG, Agarwala V, et al. Development and validation of a real-world clinicogenomic database. Journal of Clinical Oncology 2017; 35(15): 2514.

27. Birnbaum B, Nussbaum N, Seidl-Rathkopf K, et al. Model-assisted cohort selection with bias analysis for generating large-scale cohorts from the EHR for oncology research. 2020.

28. Ma X, Long L, Moon S, Adamson BJ, Baxi SS. Comparison of Population Characteristics in Real-World Clinical Oncology Databases in the US: Flatiron Health, SEER, and NPCR. 2020. doi: 10.1101/2020.03.16.20037143

29. Harrell Jr FE. Regression modeling strategies: with applications to linear models, logistic and ordinal regression, and survival analysis. Springer. 2015.

30. Platt J. Probabilistic outputs for support vector machines and comparison to regularized likelihood methods. Advances in Large Margin Classifiers 1999: 61–74.

31. Niculescu-Mizil A, Caruana R. Predicting good probabilities with supervised learning. ICML ‘05: Proceedings of the 22nd International Conference on Machine Learning 2005.

32. Goldstein M, Han X, Puli A, Perotte AJ, Ranganath R. X-CAL: Explicit Calibration for Survival Analysis. 2021.

33. Harrell Jr FE, Lee KL, Mark DB. Multivariable prognostic models: issues in developing models, evaluating assumptions and adequacy, and measuring and reducing errors. Statistics in medicine 1996; 15(4): 361–387.

34. Yu-Heng L, Chen WN, Hsu TC, Lin C, Tsao Y, Wu S. Overall survival prediction of non-small cell lung cancer by integrating microarray and clinical data with deep learning. Scientific reports 2020; 10(1): 4679.

35. Troyanskaya O, Cantor M, Sherlock G, et al. Missing value estimation methods for DNA microarrays. Bioinformatics (Oxford, England) 2001; 17(6): 520–525. doi: 10.1093/bioinformatics/17.6.520

36. Shen R, Martin A, Ni A, et al. Harnessing Clinical Sequencing Data for Survival Stratification of Patients with Metastatic Lung Adenocarcinomas. JCO precision oncology 2019; 3.

37. Thiébaut AC, Bénichou J. Choice of time-scale in Cox’s model analysis of epidemiologic cohort data: a simulation study. Statistics in medicine 2004; 23(24): 3803–3820.

38. Chiou SH, Austin MD, Qian J, Betensky RA. Transformation model estimation of survival under dependent truncation and independent censoring. Statistical methods in medical research 2019; 28(12): 3785–3798.

