## Supplemental Appendix for "Penalized regression for left-truncated and right-censored survival data"

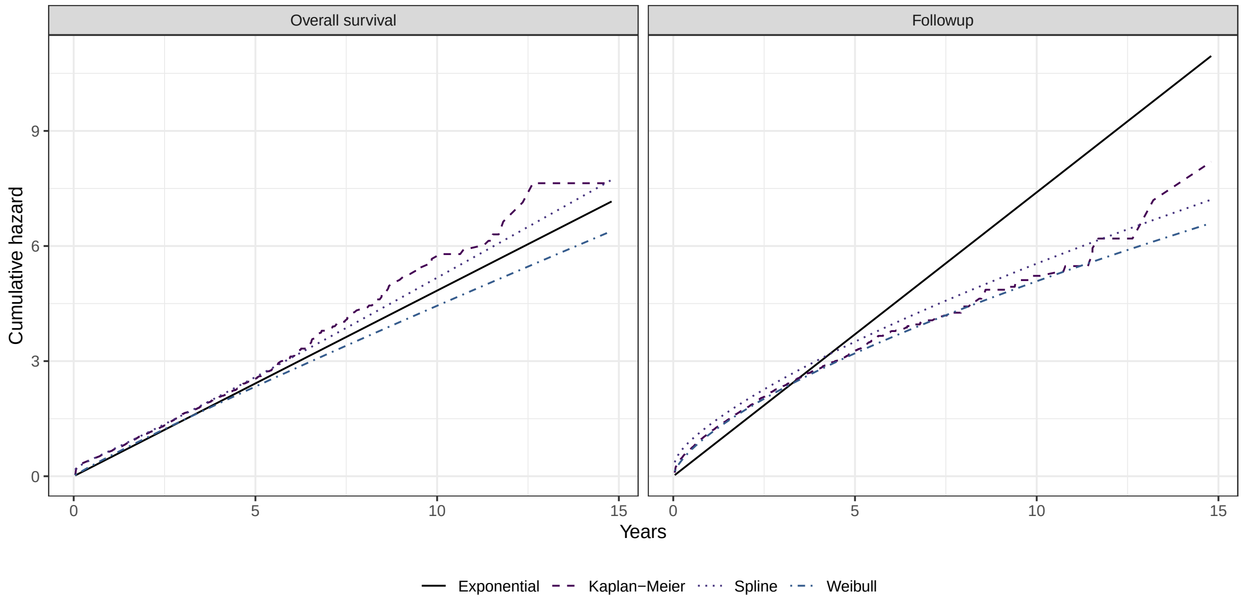


**Figure S1: Cumulative hazards among non-small cell lung cancer patients in the CGDB in models adjusting for right censoring and left truncation**

Notes: The left and right facets plot cumulative hazards for overall survival (i.e., time to death) and followup time (i.e., time to right censoring), respectively. Estimates in the rightmost plot use a "reverse Kaplan-Meier" approach whereby the meaning of an event and censoring are flipped. The spline models were fit with one internal knot at the median of the log of followup time.

*Contributed equally.

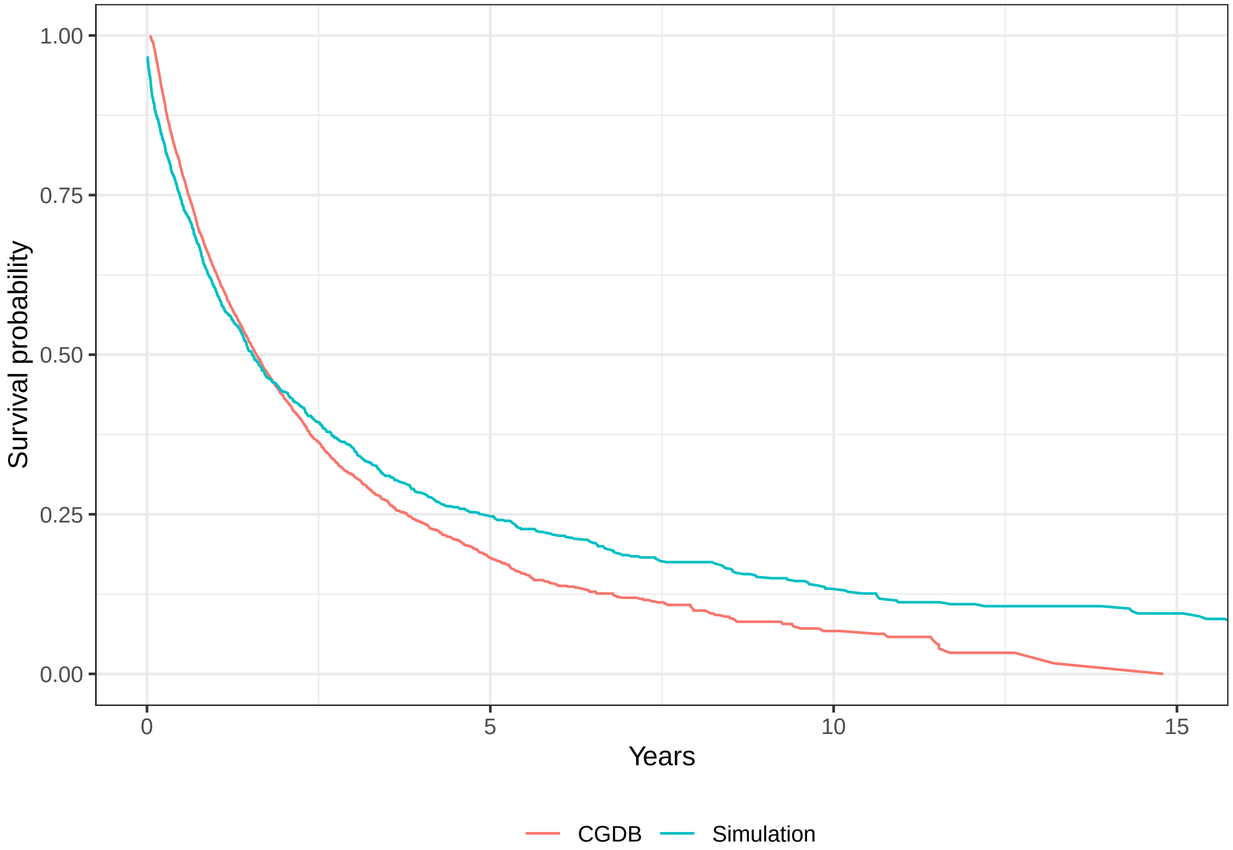


**Figure S2: Kaplan-Meier plots of overall survival from models that adjust for right censoring but not left truncation in the simulated data and CGDB**

Notes: The simulated dataset was simulated using a Weibull model including the predictors described in the main text from the CGDB. Only non-truncated patients from the simulation were used for estimation.
